# SARS-CoV-2 Seroprevalence among First Responders in Northeastern Ohio

**DOI:** 10.1101/2021.06.27.21259432

**Authors:** Xiaochun Zhang, Elie Saade, Jaime H Noguez, Christine Schmotzer

## Abstract

**Objectives:** First responders including firefighters, emergency medical technicians (EMT), paramedics, and police officers are working on the front lines to fight the COVID-19 pandemic and facing a higher risk of infection. This study assessed the seroprevalence among first responders in northeastern Ohio during May-September 2020.

**Methods:** A survey and IgG antibody test against SARS-CoV-2 were offered to University Hospitals Health System affiliated first responder departments.

**Results:** A total of 3080 first responders with diverse job assignments from more than 400 fire and police departments participated in the study. Among them, 73 (2.4%) were seropositive while only 0.8% had previously positive RT-PCR results. Asymptomatic infection accounts for 46.6% of seropositivity. By occupation, seropositive rates were highest among administration/support staff (3.8%), followed by paramedics (3.0%), EMTs (2.6%), firefighters (2.2%), and police officers (0.8%). Seroprevalence was not associated with self-reported exposure as work exposure rates were: paramedics 48.2%, firefighters 37.1%, EMTs 32.3%, police officers 7.7%, and administration/support staff 4.4%. Self-reported community exposure was strongly correlated with self-reported work exposure rate rather than seroprevalence suggesting a potential impact of risk awareness. Additionally, no significant difference was found among gender or age groups; however, black Americans have a higher positivity rate than other races although they reported lower exposure.

**Conclusions:** Despite the high work-associated exposure rate to SARS-CoV-2 infection, first responders with different roles demonstrated seroprevalence no higher than their administrative/supportive colleagues, which suggests infection control measures are effective in preventing work-related infection.

## INTRODUCTION

First responders include firefighters, emergency medical technicians (EMT), paramedics, and police officers. There are over one million first responders in the US working on the front lines fighting the COVID-19 pandemic along with healthcare workers. Due to the nature of their work, first responders face a greater risk of coming into contact with SARS-CoV-2, the causative agent of COVID-19. While personal protective equipment (PPE) and engineering controls have been used in the healthcare workplace, their effectiveness in preventing infection has not yet been gathered and thoroughly studied. Data gathered regarding exposure and infection rates among the first responders is important in providing information on the totality of undiagnosed infections and could shed light on the association between seroprevalence, sociodemographics, and occupation in the frontline critical workforce.

So far, there have been a number of publications on COVID prevalence among healthcare workers in different countries and regions ^1-5^. However, studies for first responders are scarce and limited in sample size ^6 7^. In this study, we surveyed and conducted COVID-19 antibody testing for 3080 first responders with diverse job assignments from 415 different fire and police departments in northeastern Ohio from May to September 2020. As the COVID-19 pandemic continues to evolve, information gained from this study can be helpful for the planning of future infection control strategies in this critical workforce.

## MATERIALS AND METHODS

### Study design and participants

The study was reviewed and approved by the University Hospitals Health System (UHHS) institutional review board. Participation in this research was completely voluntary. We proposed recruiting 5,000 first responders from emergency medical service, fire and police departments affiliated with UHHS in northeast Ohio. All first responders who work in the region, were older than 18 years of age and able to provide consent were invited to participate. There were no additional exclusion criteria. Work and community exposure were defined as direct contact with COVID-19 confirmed individuals at work or in the community, respectively.

### Informed consent, survey and study workflow

Participants were requested to provide informed consent, complete a survey, and get a single blood draw by venipuncture. REDCap (Research Electronic Data Capture), a secure web application, was used for administering informed consent forms electronically, managing the surveys, and storing COVID antibody results.

The study workflow is summarized in supplemental Figure 1. Department heads of UHHS-affiliated first responder departments were notified by email about the study and were asked to respond affirmatively if their departments would like to participate. A poster describing the study was then provided to interested departments to inform the potential participants about study details. Times were scheduled with the interested first responder departments to meet with potential subjects to describe the study, answer questions, allow subjects to read and complete an informed consent form and then complete the electronic survey. A single blood sample was drawn from the participant by paramedics at the first responder’s work locations or by phlebotomists at a UHHS hospital. After the blood draw, an email providing instructions for accessing results and a copy of Frequently Asked Questions about SARS-CoV-2 antibody testing was sent to participants. The SARS-CoV-2 results were provided to the participants confidentially via the secure UHHS patient portal. The employer or the supervisors of the participants did not have access to the results.

**Fig. 1.**
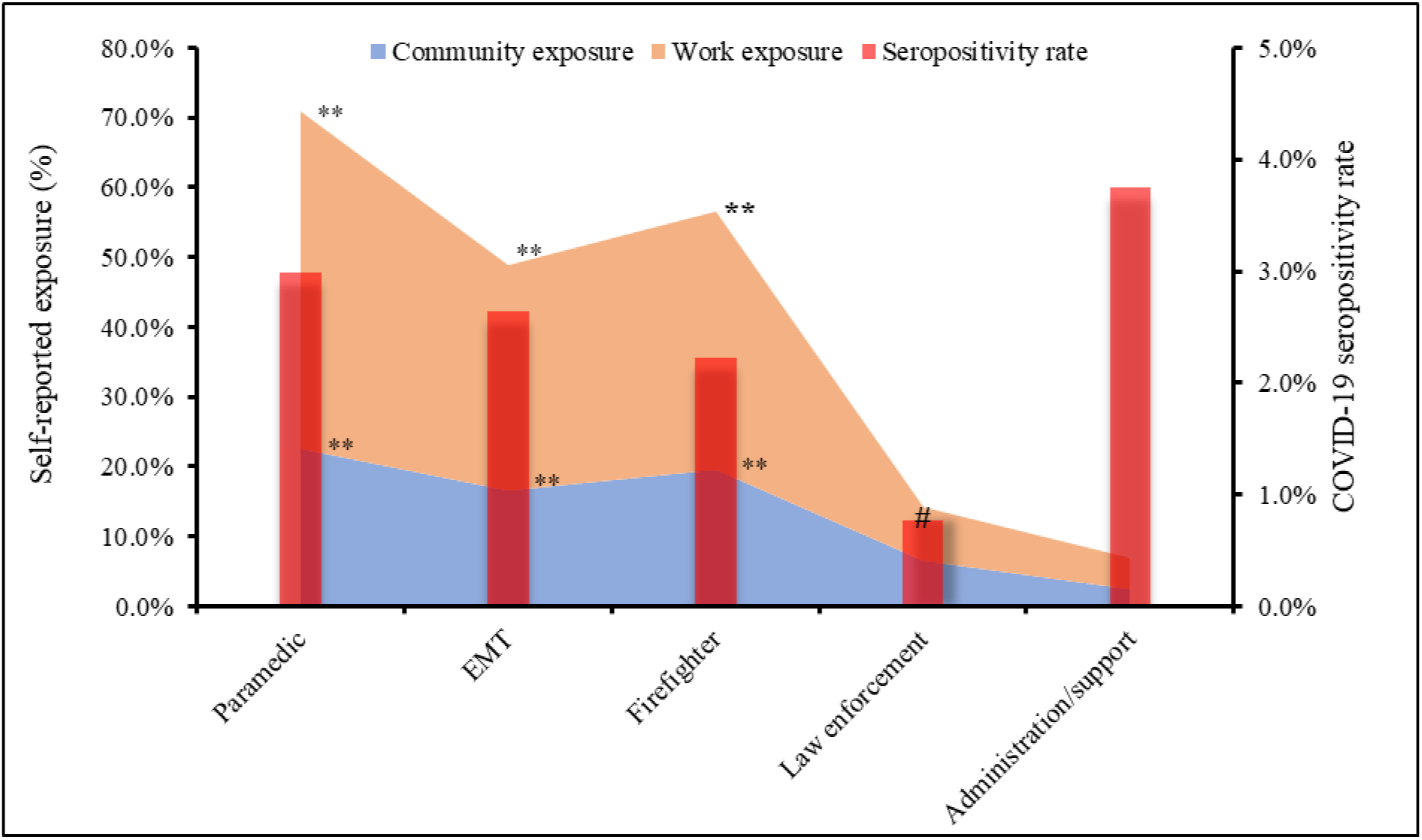
Seropositivity rate and exposure by occupation. Paramedics, EMTs, and firefighters showed significantly high exposure associated with work when compared to law enforcement and administration/support staff. Paramedics, EMTs, and firefighters also reported significantly higher community exposure. Law enforcement had the lowest seropositivity rate compare to other occupations. **: Exposure rate p<0.01. #: Seropositive rate p<0.05.

### Specimen collection and processing

A minimum of 1 ml of blood was collected into a serum separator tube by venipuncture. Serum samples were allowed to clot adequately before centrifugation and then centrifuged at 3,000 rpm for 6 minutes. After centrifugation, samples were stored at room temperature (15 - 30°C) for 2 days or 7 days at 2 - 8°C before testing.

### SARS-CoV-2 antibody testing

The antibody testing was conducted at the University Hospitals Cleveland Medical Center core laboratory which is certified under the Clinical Laboratory Improvement Amendments (CLIA) of 1988, 42 U.S.C 263a, to perform high complexity testing. The testing was performed using the Abbott SARS-CoV-2 IgG assay, a chemiluminescent microparticle immunoassay on Architect i1000SR analyzers for detecting IgG antibodies against the nucleocapsid protein of SARS-CoV-2. This assay has been approved by the FDA for use under an Emergency Use Authorization (EUA). Test results were reported qualitatively as negative or positive based on index values using a cutoff of ≥1.4.

### Statistical analysis

Statistical analyses were performed using SigmaPlot software. The categorical variables were presented as percentage and the Chi-square test or Fisher’s exact test was used to determine significance. The SigmaPlot software automatically analyzes data for its suitability for Chi-Square or Fisher’s exact test and suggests the appropriate test based on the sample size of each group. Possible associations between exposures and seroprevalence were assessed using Relative Risk (RR). The continuous variables were expressed as mean and SD and/or median with quartiles. The difference between two groups was compared using the Student t test or Mann-Whitney Rank Sum Test depending on whether the Normality Test (Shapiro-Wilk) passed or failed. The difference among three or more groups was compared using the One-way ANOVA or Kruskal-Wallis One-way ANOVA on ranks depending on the result of the Normality Test. Spearman Rank Order Correlation was used to assess the correlation between variables such as seroprevalence and exposure to COVID-19. A p value <0.05 was regarded as statistically significant.

## RESULTS

### Seroprevalence and Self-reported exposure by occupation

A total of 3,275 first responders from 415 fire and police departments in Ohio consented to the study from May 22^nd^ to September 15^th^, 2020. Among the participants, 195 (6.0%) did not get blood drawn for antibody testing and were excluded from data analysis. The 3,080 participants who completed the anti-SARS-CoV-2 antibody test were included. These participants had diverse occupations and job assignments within the first responder departments. Participants’ occupational constituents are summarized in supplemental Figure 2. Firefighters, paramedics, police officers, and EMTs were among the major occupations and accounted for 92.2% of the participants. Additionally, administration and support staff working in the fire and law enforcement departments were also included in the study, and they made up 5.2% of the participants. They were office or field staff, such as ambulette drivers and sanitation workers. Administration and support staff showed no significant difference in seroprevalence or exposure and were combined due to the small sample size.

**Fig. 2.**
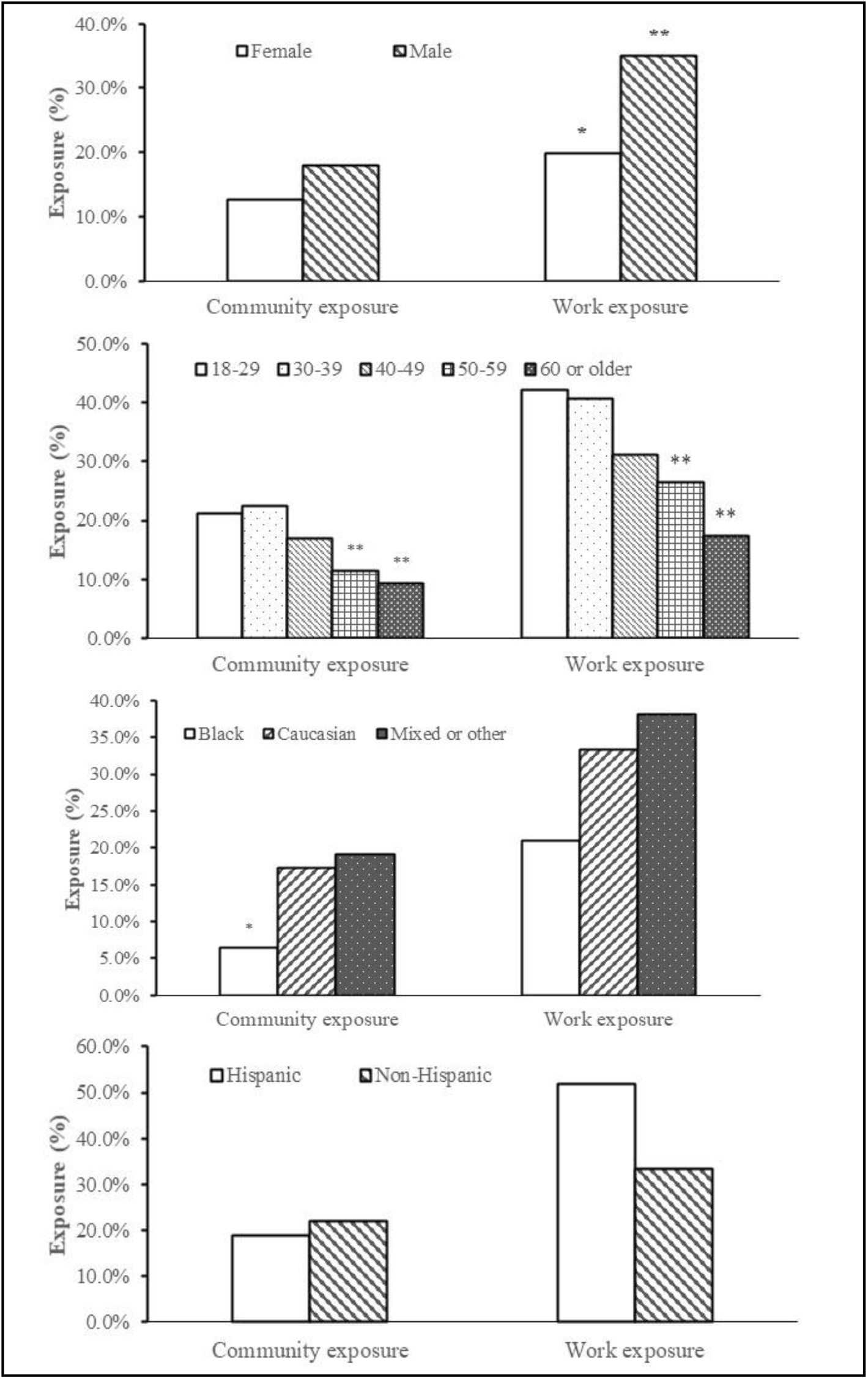
Participants self-reported exposure at work and in the community by age, gender, race, and ethnicity (n=3080). **:p<0.01. *p<0.05. Male participants had a significantly higher self-reported exposure rate associated with work than female participants. Younger participants (≤50) showed a significantly higher exposure rate than older participants (>50) both at work or in the community. Africa American participants reported a notably lower community exposure rate. Work exposure and community exposure rate were strongly correlated (Correlation Coefficient=0.81; p<0.01) despite gender, age, or race.

Seropositivity rate, work exposure, and community exposure among different occupation groups were summarized in Figure 1. By occupation, the highest level of reported work-related exposure was found among paramedics (48.2%) followed by firefighters (37.1%), EMTs 32.3%, and the lowest among police officers (7.7%) and administration/support staff (4.4%).

A total of 73 participants were positive, giving an overall seropositivity rate of 2.4%. Seroprevalence was highest among participants holding administration/support positions (3.8%) followed by paramedics (3.0%) and lowest among law enforcement/police officers (0.8%). Compared to paramedics, police officer’s RR was 0.26 (95%CI: 0.08-0.85; P<0.05). Further analysis showed no correlation between the work exposure rate and seroprevalence among different occupation groups (P>0.05).

The self-reported community exposure was also significantly different by occupation (p<0.01). There were 22.6% paramedics, 19.5% firefighters, and 16.5% EMTs who reported community exposure to COVID-19, while only 6.6% of law enforcement/police officers and 2.5% of administration/support staff reported community exposure. No correlation was found between community exposure rate and seropositivity rate. Interestingly, a strong correlation between work exposure rate and community exposure rate was observed across occupation groups (Correlation Coefficient=1.00; p<0.01).

### Seroprevalence and self-reported exposure by age, gender and race

Seropositive rates by age, gender, and race were summarized in Table 1. Female participants had a numerically higher seropositivity rate than males but the difference did not reach statistical significance. Seroprevalence was numerically higher among younger participants (18-30 years old) than other age groups, but the difference was not statistically significant. Black participants had a statistically significant higher positivity rate than other races. The RR was 3.63 (95% confidence interval (CI): 1.52 to 8.70; p=0.01) when compared with Caucasian participants.

**Table 1.**
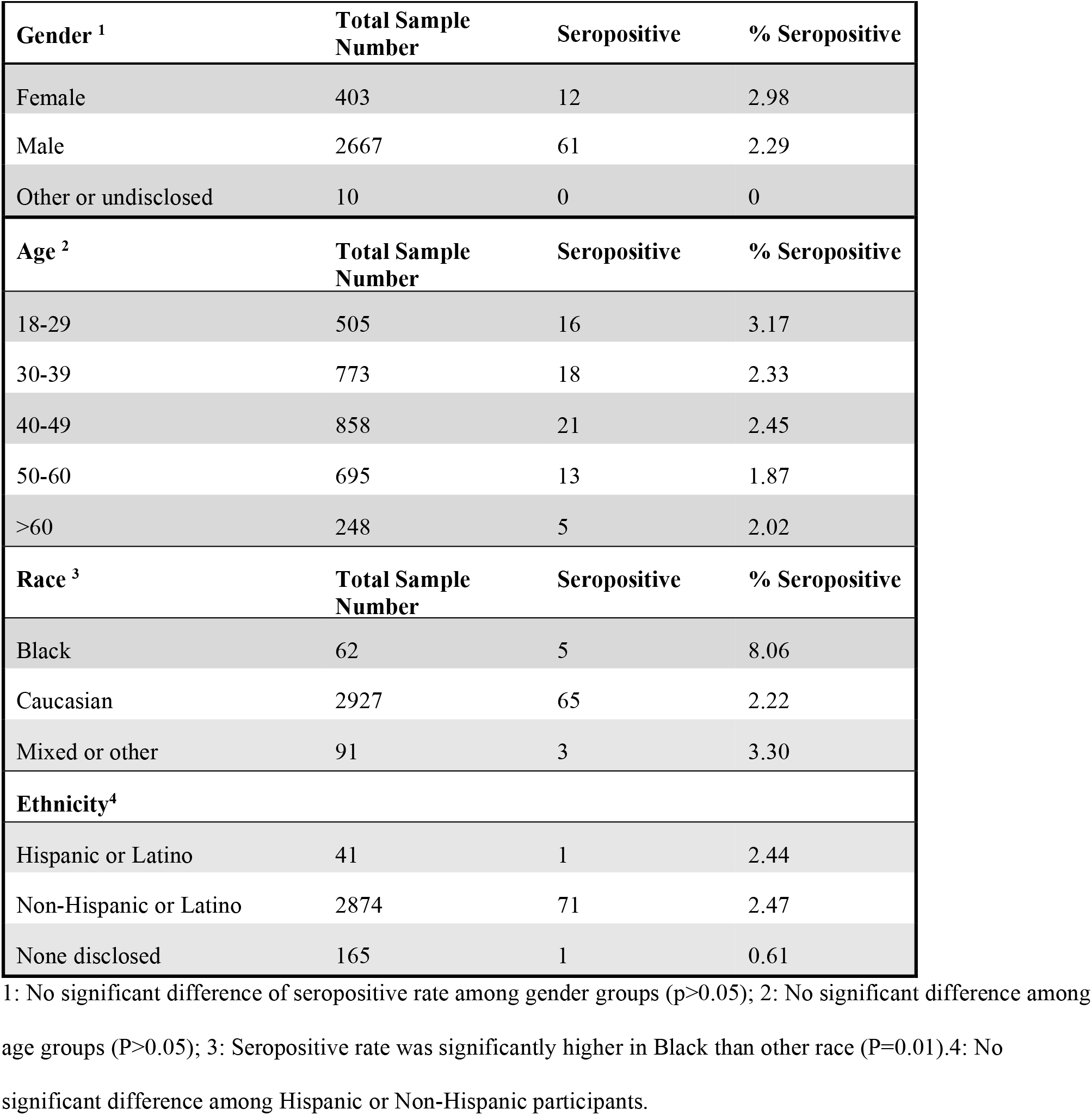
**Sociodemographic characteristics of study participants and COVID-19 seropositivity rate (n=3080)**

Self-reported exposure rates by gender, age, race, and ethnicity are shown in Figure 2. Male participants reported a similar community exposure rate but significantly higher work exposure than females. The work-related exposure was 35.1% among male participants and 19.9% among female participants. Black Americans reported significantly lower community exposure than Caucasian participants (6.5% versus 17.3%; p<0.01). Older participants (≥ 50-year-old) had lower work and community exposure compared to younger participants. Work exposure rate was positively correlated with community exposure rate (Correlation Coefficient=0.81; p<0.01), independent of gender, age, or race. However, neither the self-reported work exposure rate nor the community exposure rate correlated with the seropositivity rate. As shown in Figure 3, the seropositivity rate was not significantly different between the participants with and without self-reported exposure.

**Fig. 3.**
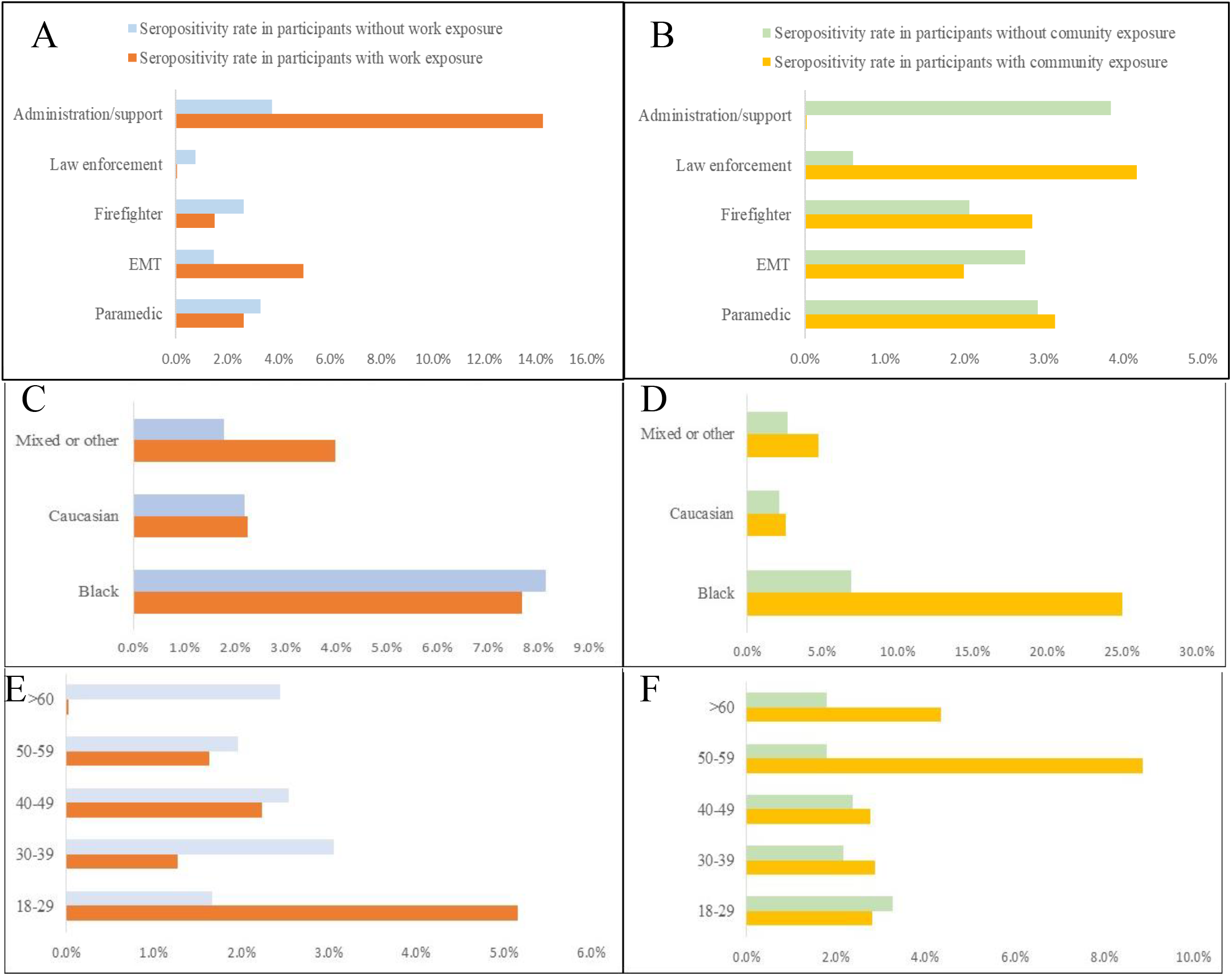
Self-reported exposure and SARS-CoV-2 seropositivity rate by occupation, race, and age (n=3080). The seropositivity rate between the participants with and without exposure was not statistically significant despite occupation, race, or age. Seropositivity rate between participants with and without work exposure by occupation (A), race (C), and age (E). Seropositivity rate between participants with and without community exposure by occupation (B), race (D), and age (F). Some subgroups showed a numerically higher seropositivity in exposed participants. However, the difference was not statistically significant (p >0.05).

### History of COVID RT-PCR test in study participants

Among the 3,080 participants, 267 people reported a history of being tested by the COVID RT-PCR test. The RT-PCR testing rate among the participants with symptoms was 29.3%. For the participants that were tested, 26 of them (0.8%) reported a positive RT-PCR result and 17 (65.4%) of them were also seropositive.

### Antibody index values

As shown in Figure 4, the antibody index of seropositive participants was dramatically higher than those with a negative result. The median antibody index was 5.8, 4.5, and 3.9 for the seropositive participant with negative PCR result, positive historical PCR result, or without a PCR test, respectively. The difference associated with PCR testing status was not statistically significant. The median index was 0.03, 0.65, and 0.03 for seronegative participants with negative PCR results, positive PCR results, or without a PCR test, respectively. The median index of PCR positive but antibody negative participants was significantly higher than others (p<0.01). Their antibodies were tested 39-142 days after the positive PCR test. Without longitudinal data, it could not be determined if the elevated but below cutoff index values represent waning antibody levels over time, less robust initial humoral immune response or both. Additionally, the antibody index was not significantly different between the seropositive participants with or without symptoms.

**Fig. 4.**
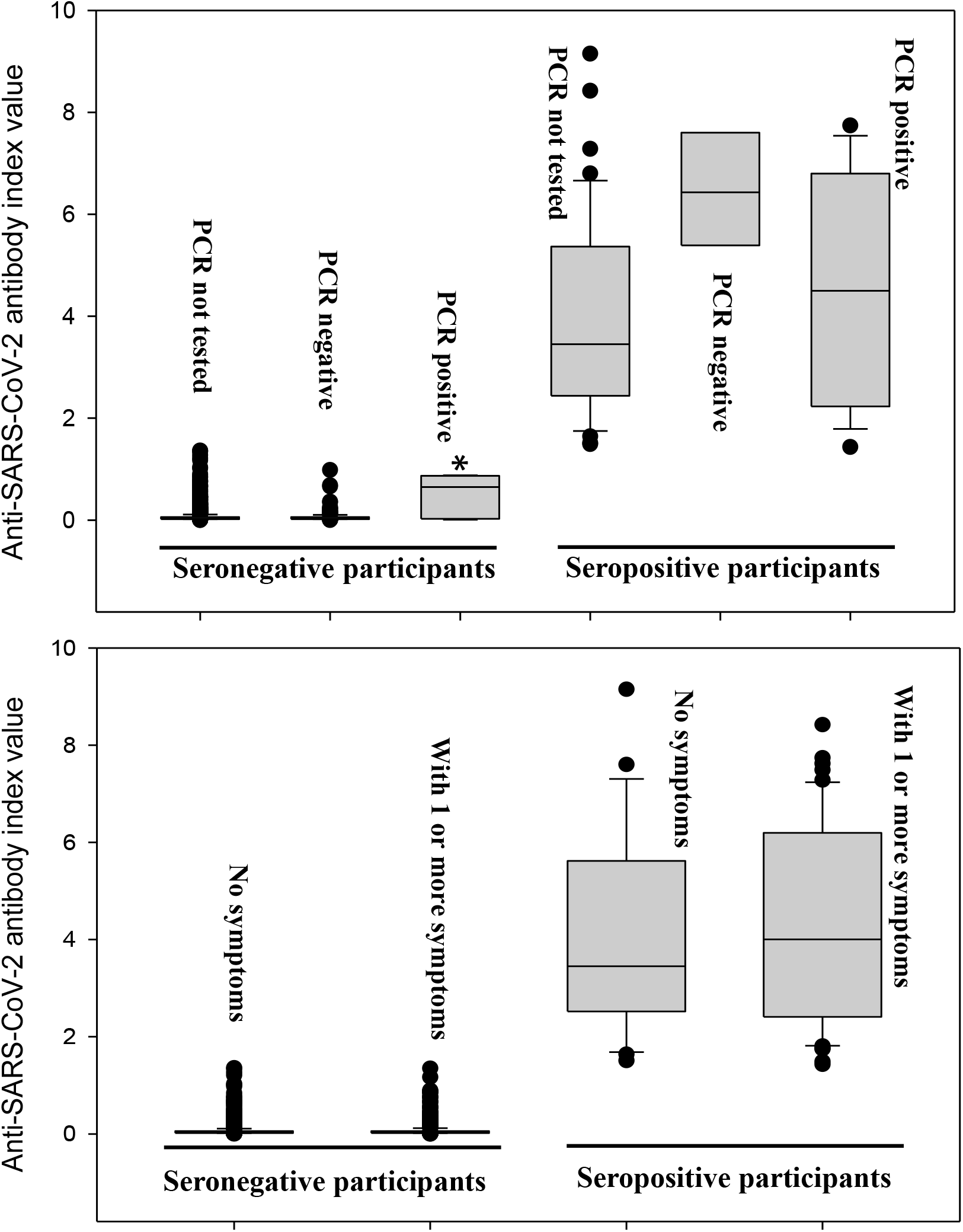
Anti-SARS-CoV-2 IgG antibody index values by PCR testing status and COVID related symptoms. Top: Index value in PCR test positive, negative, or not performed individuals. *p<0.05. Among antibody-negative participants, the ones with a previous positive PCR result showed a much higher median index (0.65) than the PCR negative (0.03) or not tested seronegative individuals (0.03). No significant difference in index values was found among the seropositive participant neglecting PCR testing status. Bottom: Median antibody index was slightly higher among seropositive participants who reported one or more COVID-related symptoms but not statically significant.

### Self-reported symptoms and correlation with seroprevalence

Among the seropositive participants, 53.4% reported one or more symptoms and 46.6% were asymptomatic. Meanwhile, 28.3% of seronegative participants also reported diverse symptoms. Supplemental Table 1 compared the frequency and odds ratio of common symptoms between the COVID seropositive and seronegative participants. Loss of smell/taste was a common and relatively specific symptom for COVID with an odds ratio of 20. Shortness of breath, fever, muscle ache, and diarrhea were 2 to 3 times more frequent in seropositive than seronegative participants. Cough, sore throat, and runny/stuffed nose were common and presented at a similar rate in seropositive and seronegative participants. Conjunctivitis or red eye was rare (<5%) and had no strong correlation with COVID seropositivity in this population.

## DISCUSSION

There are a number of epidemiologic studies which assessed SARS-CoV-2 infection rates and risk factors in healthcare workers worldwide, but knowledge about exposure and infection among first responders is very limited. This study provides a thorough evaluation of COVID-19 seroprevalence and exposure in first responders with 3080 participants across diverse job duties during the first and second wave of the pandemic in the U.S.

Despite their limitations, serological tests are important tools for assessing SARS-CoV-2 infection and potential immunity. Fifty-six antibody assays have received emergency use authorization (EUA) from the Food and Drug Administration (FDA) by December 20, 2020. In general, lab-based assays, especially chemiluminescent microparticle immunoassay assay performed on fully automated analyzers, have higher specificity compared to rapid tests (later flow immunoassay) according to the performance characteristics shown on the FDA webpage (https://www.fda.gov/medical-devices/coronavirus-disease-2019-covid-19-emergency-use-authorizations-medical-devices/eua-authorized-serology-test-performance). Assay specificity is critical in reducing false-positive rates for large scale survey in low prevalence population. Per the manufacturer’s package insert, the assay used in this study was estimated to have 100% sensitivity (>14 days post-symptom onset) and 99.6% specificity. An independent evaluation study by Bryan et al also confirmed excellent specificity (99.9%) of this assay ^8^. The estimated Positive Predictive Value and Negative Predictive Value at a prevalence of 5% were calculated to be 93.4% and 100%, respectively.

By occupation, paramedics, EMTs, and firefighters showed similar seropositivity rates around 2-3%. The positivity rates are slightly lower than administrative or support staff. Analysis of self-reported exposure confirmed the high-risk nature of these occupations had significantly higher work-related COVID-19 exposure. The average exposure rate was 39.2% and about 5 times higher than the occupations that do not need to encounter known or suspected COVID-19 patients such as administrative/support staff. Although the study survey did not assess PPE usage, proper PPE wearing is part of the required training and workplace policy at UHHS and its affiliated facilities. Taken together, the results suggest the effectiveness of the safety measures and PPE in protecting from work-related infection. This is also supported by comparing COVID prevalence in the general population. The Ohio statewide prevalence of current and past COVID-19 infection was estimated to be 0.9% and 1.5% respectively by the end of July 2020 (https://coronavirus.ohio.gov/static/dashboards/prevalence-covid19-ohioadults.pdf). These data were from a study performed by the Ohio Department of Health and the Ohio State University during July 9-28th on 727 Ohio adults using PCR and antibody tests in combination with mathematic models. In our study, 2,413 participants had been tested by the end of July with a positive rate was 2.2%. Furthermore, we compared our findings with similar studies. So far, two studies reported serosurvey results among first responders. One study performed in mid-April by Caban-Martinez et al. reported an 8.9% seropositive rate among 203 firefighters/paramedics from a single fire department in Florida ^6^. However, the study used a rapid antibody test with 90.6% specificity and the positivity rate might be overestimated. The second study was a large-scale study including both first responders and healthcare workers conducted by the Center for Disease Control and Prevention (CDC) during May-June, 2020 in the Detroit Metropolitan Area and Michigan which used a fully automated chemiluminescent immunoassay but with a different antigen target ^9^. The study, however, did not further differentiate paramedics, EMTs, or firefighters. Among the about 2000 first responders, the seropositive rate was 5.2%. Similar to our results, the first responder’s seropositive rate was lower than the administration/clerk (8.0%).

Among the different occupations, police officers had the lowest seroprevalence (0.8%). This finding is also consistent with the CDC study which reported low seroprevalence in police/corrections officers as compared to EMS, firefighters, and healthcare workers ^9^. Police/law enforcement officers in our study reported similar levels of work and community exposure to administrative/support staff.

Our results also showed a trend of high seroprevalence among Black-American participants ^10-12^. The self-reported exposure at work and community were much lower than Caucasian counterparts. The notable lower exposure, unlikely to reflect the actual risk, may rather suggest a potentially low testing rate and a high rate of asymptomatic infection among the community.

The finding of a significantly higher seropositivity rate compared to PCR positivity rate (2.4% vs. 0.8%, p<0.01) is consistent with other serology surveys ^10 13 14^. Additionally, a strong correlation (r = 0.83; P<0.01) between the seropositivity rate and PCR positivity rate was found across occupations, race, and gender. The seropositivity rate was estimated to be 3.2±0.9 times of the PCR positivity rate. This estimation is similar to the 2.5 ratio by the CDC study which reported 6.9% seroprevalence among the 16,403 healthcare workers and first responders with 2.7% having a history of a positive RT-PCR test ^9^. There are multiple potential underlying reasons for the higher seroprevalence versus PCR positive rate. First, nearly half of the seropositive participants (46.6%) reported no COVID-related symptoms. These asymptomatic individuals are unlikely to request PCR testing. Second, the PCR testing rates among the symptomatic individuals were as low as 30% among first responders. Finally, 7 PCR negative participants were seropositive. Given the high specificity of the antibody assay and high SARS-CoV-2 index values of those samples, these are likely to be true positive antibody results missed by the RT-PCR test, which accounts for 9.6% of the seropositive participants.

One of the limitations of this study is that 9 out of 26 (34.6%) participants with previous positive COVID PCR test were seronegative, indicating that the seropositivity rates among first responders were potentially underestimated in this study. The underestimation may be attributed to attenuated antibody concentration over time, a weak humoral immune response associated with mild symptoms, or production of antibodies not targeting the N protein. Additionally, self-reported exposure was not further categorized by duration, distance, or PPE usage. The survey questions were designed to balance between information collection and time constraints. Some valuable details were challenging to collect from the busy first responders during a pandemic.

In conclusion, the study showed first responders had a similar infection rate to their coworkers whose duties do not involve direct patient contact. The results indicate the effectiveness of infection control measures implemented and compliance of first responders in following safety procedures. Additionally, the high rate of asymptomatic infection and miscorrelation between seropositivity rate and self-reported exposure underscores the risk posed by undiagnosed infections.

## Data Availability

Data available on request from the authors.

## ACKNOWLEDGMENTS

We would like to thank all of the participants for their contribution in expanding our knowledge about COVID-19 epidemiology. We would like to thank individuals within the following entities whose efforts made this study possible: the UH EMS Training and Disaster Preparedness Institute, Eileen Terrell and the Harrington Heart and Vascular Institute Research staff, University Hospitals Laboratories staff, and the RedCap team. This study was supported by the generosity of donors.

---

**Supplemental Fig. 1.**
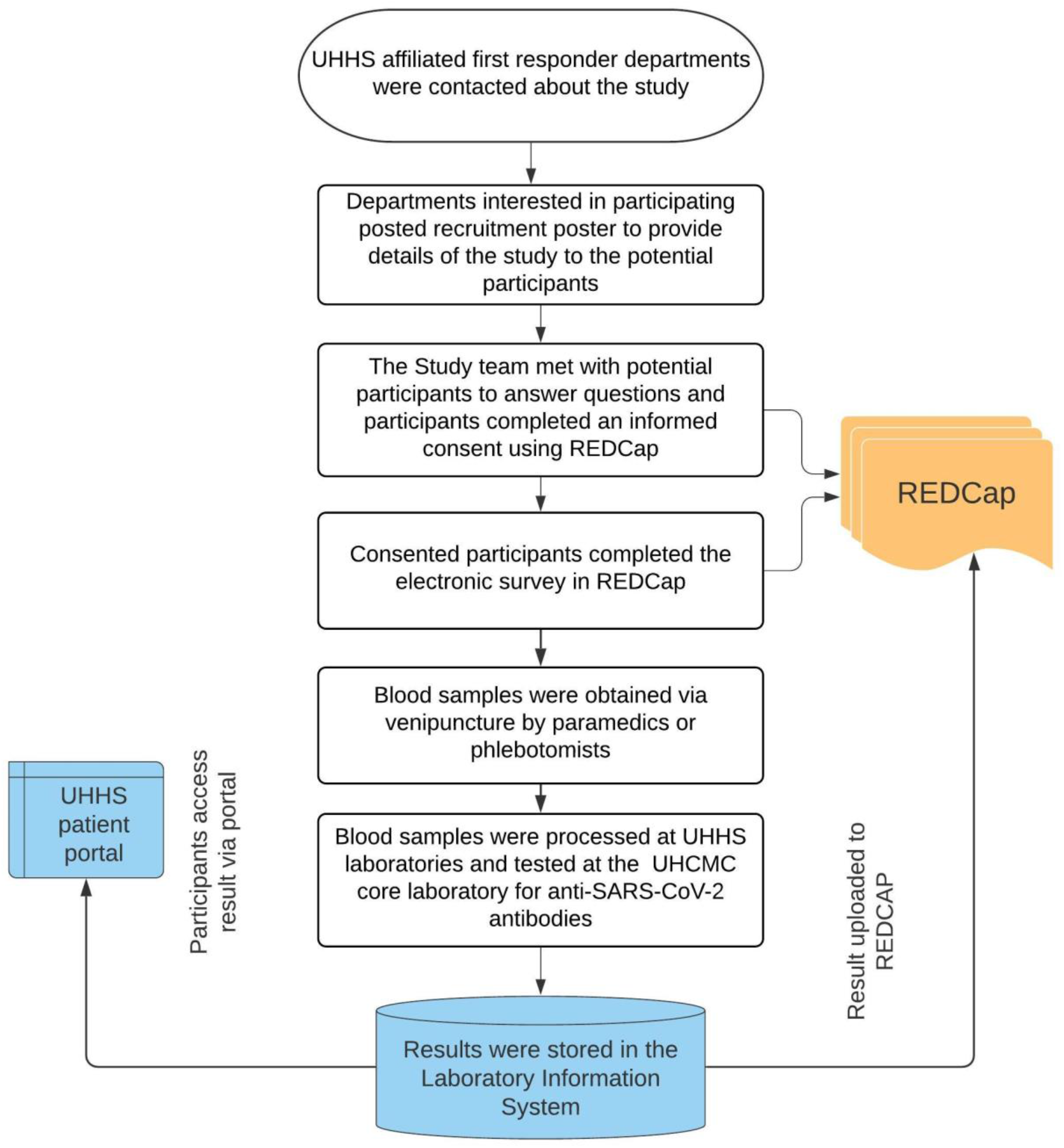
Study flowchart. The flowchart summarized the overall study process including recruitment, consent, sample processing, and data flow. UHHS: University Hospitals Health System. UHCMC: University Hospitals Cleveland Medical Center. REDCap: Research Electronic Data Capture, a secure web application for research surveys and databases.

**Supplemental Fig. 2.**
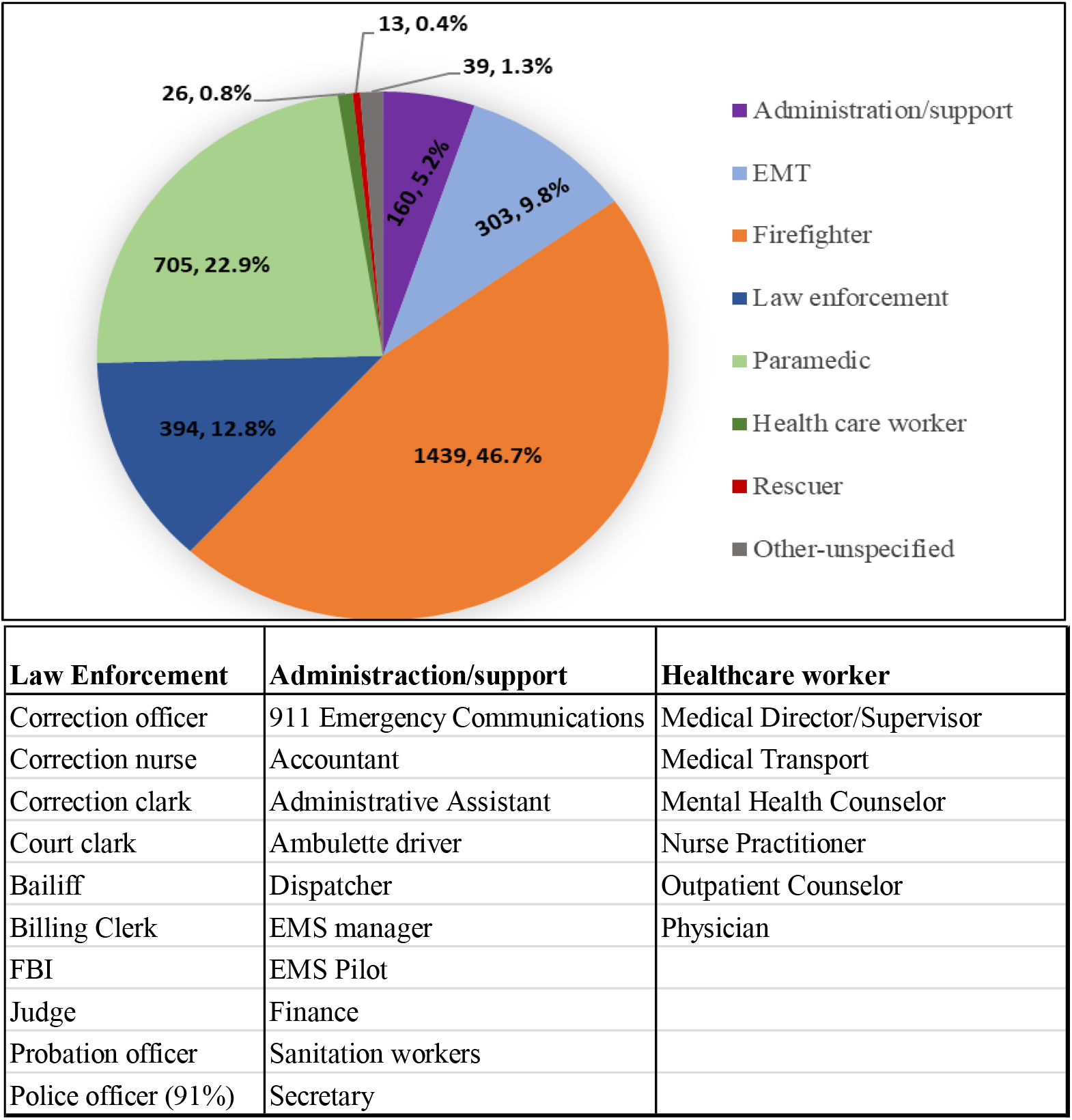
Occupation and job assignment of study participants (n=3080). Top: Participants’ occupation categories. Bottom: Job assignments included in the occupation categories. In the law enforcement category, 91% (358 out of 394) were police officers.

**Supplemental Table 1.** **Self-reported symptoms since February 1^st^ 2020 and odds ratio between COVID-19 seropositive and seronegative participants (n=3080)**

|  |  |  |  |  |
| --- | --- | --- | --- | --- |
| Symptoms | % in seropositive participants | % in seronegative participants | Odds ratio | p value |
| Loss of taste or smell | 26.0% | 1.7% | 19.99 | <0.01 |
| Shortness of breath | 17.8% | 7.4% | 2.70 | <0.01 |
| Fever | 21.9% | 9.7% | 2.62 | <0.01 |
| Muscle aches | 24.7% | 12.9% | 2.20 | <0.01 |
| Diarrhea | 15.1% | 7.7% | 2.12 | <0.05 |
| Conjunctivitis or red eye | 4.1% | 1.5% | 2.82 | >0.05 |
| Sore throat | 19.2% | 13.0% | 1.59 | >0.05 |
| Cough | 27.4% | 19.5% | 1.56 | >0.05 |
| Runny or stuffed nose | 21.9% | 19.1% | 1.18 | >0.05 |

